# Clinical presentation and in-hospital outcomes of older patients hospitalized with COVID-19 in Montreal, Canada: a retrospective review

**DOI:** 10.1101/2021.02.27.21252596

**Authors:** Sandrine Couture, Marc-Antoine Lepage, Claire Godard-Sebillotte, Nadia Sourial, Catherine Talbot-Hamon, Richard Kremer, Ami Grunbaum

## Abstract

**Background:** Older adults are more vulnerable to severe infection and mortality due to COVID-19. They often have atypical presentations of the disease without respiratory symptoms, which makes early diagnosis clinically challenging. We aimed to compare the baseline characteristics, presentation, and disease course of older and younger patients hospitalized with COVID-19.

**Methods:** The charts of 429 consecutive patients hospitalized in Montreal, Canada, with PCR-confirmed COVID-19 were retrospectively reviewed. Baseline health, presentation, in-hospital complications, and outcomes were recorded. Desegregation by age was performed to compare older (≥70) versus younger (<70) individuals.

**Results:** Older patients presented with more comorbidities compared to younger patients as captured by the Charlson Comorbidity Index (mean 6 vs 2), including higher rates of cardiovascular, cerebrovascular, chronic obstructive pulmonary, and chronic kidney disease. Older patients were less likely than younger patients to present with cough (27% vs 47%) or dyspnea (33% vs 48%). Fifty-two (52%) had no respiratory symptoms on presentation compared to 32% in the younger group (p<0.001); however, they were more likely to present with geriatric syndromes such as delirium (29% vs 7%), functional decline (14% vs 0.6%), or falls (15% vs 5%). Twelve (12%) of older patients presented with a geriatric syndrome as their sole symptom compared to 3% in the younger group (p=0.002). Older adults were more likely to develop acute kidney injury (35% vs 22%), malnutrition (9% vs 4%), delirium (29% vs 17%) and hypernatremia (32% vs 17%). They had higher in-hospital mortality (33% vs 13%, p<0.001).

**Discussion:** Older adults presenting to hospital with COVID-19 commonly have no respiratory symptoms and can present with only a geriatric syndrome. A new geriatric syndrome in an older person should trigger isolation and evaluation for COVID-19. Furthermore, older adults are particularly vulnerable to complications related to dehydration, warranting early initiation of multidisciplinary care.

## INTRODUCTION

Older adults are amongst the most vulnerable to severe infection and mortality due to COVID-19 (1-3). In Canada, as of January 2021, adults above age 70 accounted for 55% of COVID-19 hospitalizations and 89% of deaths (4), while representing only 12% of the population (5). There is growing evidence that older adults are more likely to have atypical presentations of COVID-19 with lack of fever, cough, or dyspnea (6-8). Emerging reports suggest that COVID-19 may present as geriatric syndromes including falls and delirium in this population (8, 9). The nuances of the clinical presentation of older patients with COVID-19 has critical implications for the early diagnosis, timely isolation and infection control. An improved understanding of the disease course in older adults is crucial for enabling optimal care and prognostication in this high-risk population. To date, Canadian data is lacking with regards to clinical presentation, demographics, comorbidities and disease course of older adults hospitalized with COVID-19. The aims of this study are 1) to describe the clinical characteristics and outcomes of older persons (≥70) hospitalized with COVID-19 at a tertiary hospital in Montreal during the first wave, 2) to determine whether older adults exhibit different presentations compared to younger patients (<70) and 3) to determine whether older adults have different in-hospital course compared to younger patients. We hypothesized that non-respiratory presentations and geriatric syndromes would be more common in older adults, and that they would have higher mortality compared to younger patients.

## METHODS

We conducted a retrospective review of electronic medical records of all patients age 18 and above with laboratory-confirmed COVID-19 admitted to the McGill University Health Centre (MUHC), a Montreal tertiary care hospital between March 26^th^ and July 16^th^, 2020. A confirmed case of COVID-19 was defined as a positive result of a real-time reverse-transcriptase–polymerase-chain-reaction (RT-PCR) assay of a nasal swab specimen.

### Data Collection

For each patient, the following data were extracted from electronic medical records: baseline demographic information, living situation, pre-existing medical conditions, home medications, clinical presentation, and disease course pre-hospitalization. The Charlson Comorbidity Index (10) was calculated using these baseline patient characteristics to capture the degree and severity of comorbidities in our cohort. We extracted laboratory and imaging findings at the time of admission, as well as complications, need for intensive care or ventilatory support, discharge destination or in-hospital death until July 16^th^, 2020, irrespective of the date of admission.

### Definitions

Patients ≥70 were classified as older patients; those <70 years old were classified as younger patients. We defined non-respiratory presentations as absence of cough, dyspnea, hemoptysis, sputum production, nasal congestion, and sore throat. In keeping with the American Geriatrics Society’s recommendations to define geriatric syndromes as common conditions in older adults having significant consequences for functioning and quality of life (11), we considered falls, delirium and functional decline.

### Statistical Analysis

Data was analyzed using IBM SPSS Statistics for Mac version 27.0 (SPSS Inc. Chicago, IL). Continuous variables are presented as means and standard deviations. Categorical variables are summarized as counts and percentages. We used the χ^2^ test to test our hypotheses that older patients would have more atypical presentations with lack of respiratory symptoms, geriatric syndromes, and higher mortality. All statistical tests were two-sided, and the differences were considered statistically significant at p < 0.05.

### Ethics Approval

MUHC institutional review board approval was obtained for data collection. All patient information was deidentified with respects to names, admission date and date of discharge or death.

## RESULTS

### Population Characteristics and Medical Status at Baseline

In total, 429 patients were hospitalized with PCR-confirmed COVID-19 at the MUHC between March 26^th^ and July 16^th^, 2020. Socio-demographic and baseline health characteristics of our cohort are presented in Table 1. Sixty-one percent (61%) were aged 70 and above. Average age was 83 years old in the older group (standard deviation 7.31) and 55 in the younger group (standard deviation 11.71). Forty-nine percent (49%) of patients in the older group and 37% in the younger group were women.

**Table 1.**
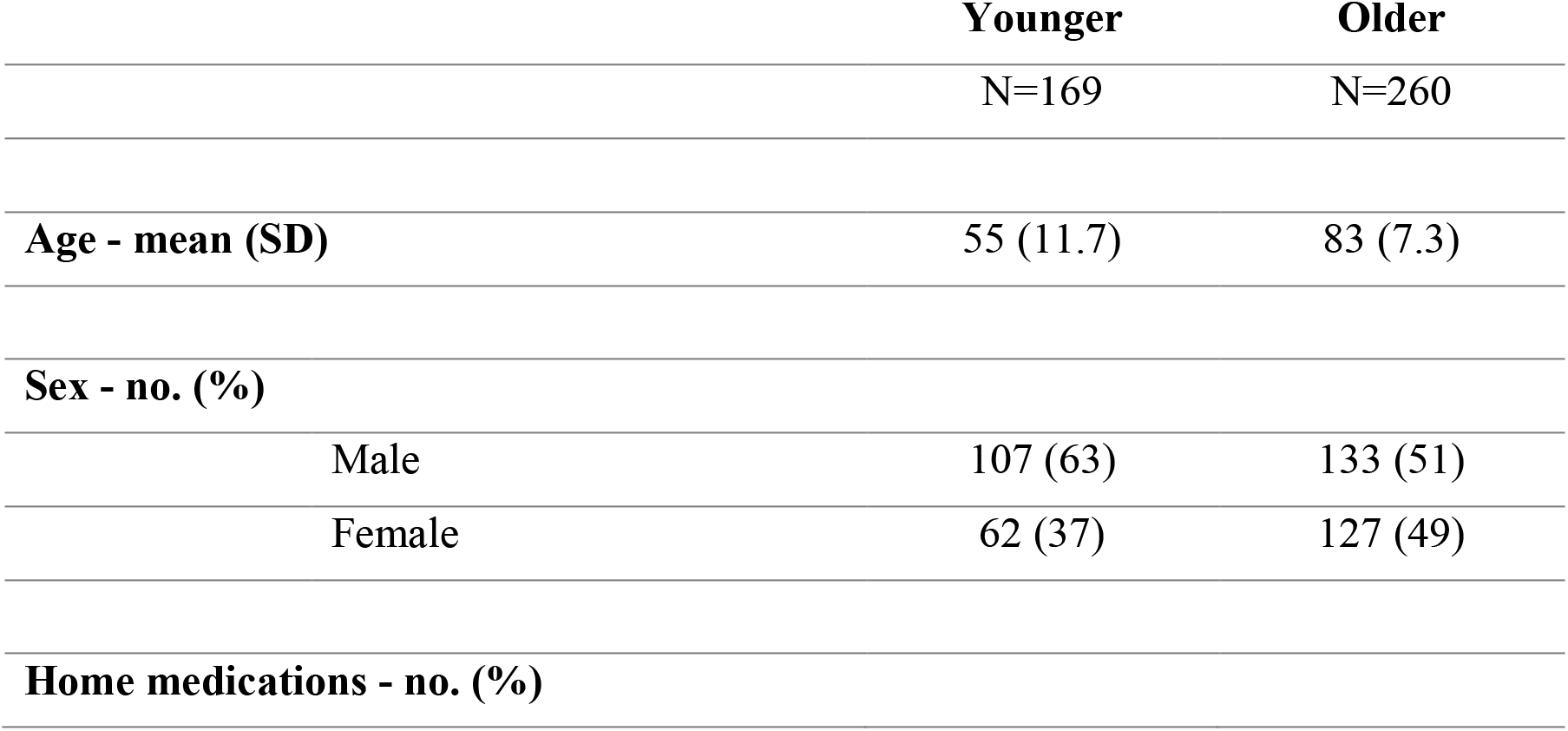
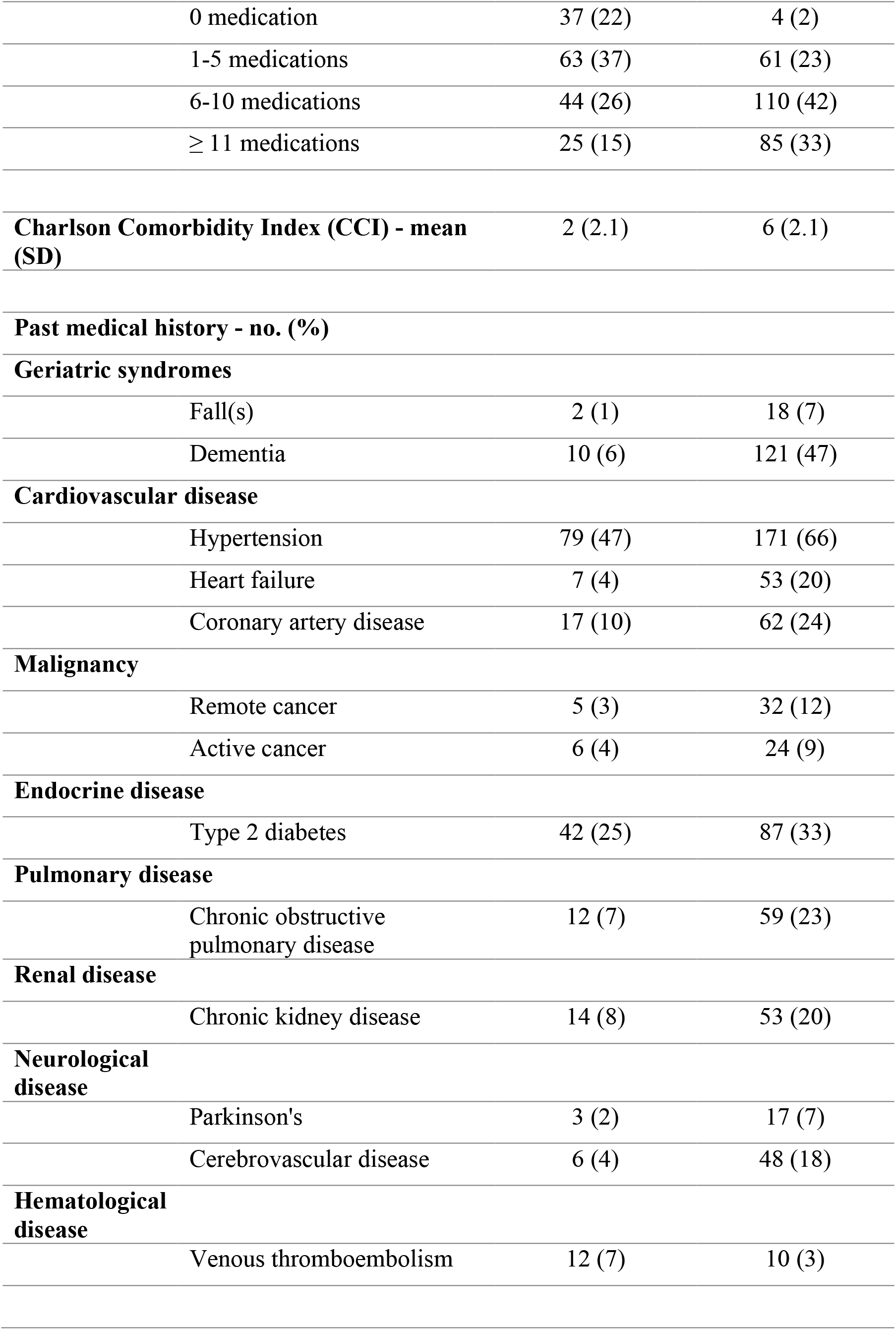
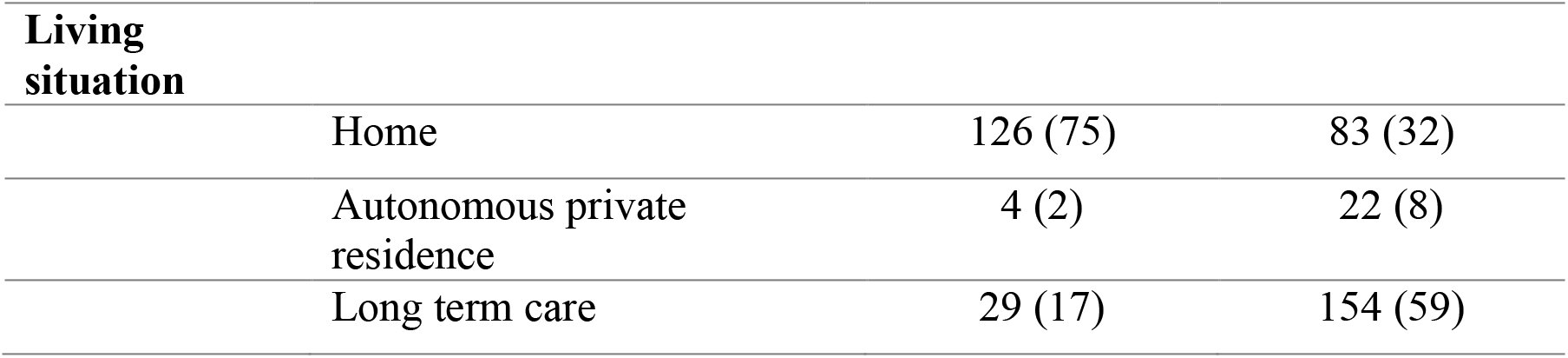
Population Characteristics.

Older patients had more pre-existing medical conditions at baseline as captured by a higher mean Charlson Comorbidity Index of 6 compared to 2 in the younger group. These comorbidities included higher proportions of hypertension, coronary artery disease, heart failure, cerebrovascular disease, chronic obstructive pulmonary disease, chronic kidney disease (CKD), major neurocognitive disorder, and active cancers.

This higher proportion of comorbidities in the older group was reflected by a greater number of home medications compared to the younger group. Seventy-five percent (75%) of patients in the older group had polypharmacy (>5 medications) compared to 41% in the younger group.

Fifty-nine percent (59%) of older patients originated from long-term care facilities and 32% were living at home in the community.

### Clinical Presentation and Laboratory Abnormalities

Older patients were less likely to present to hospital with respiratory symptoms of cough (27% vs 47%) and dyspnea (33% vs 48%) compared to their younger counterparts. Fifty-two percent (52%) of older patients presented without any respiratory symptoms compared to 32% of younger patients (p<0.001).

Older adults were more likely than younger patients to present with delirium (29% vs 7%), functional decline (14% vs 1%), and falls (15% vs 5%). Twelve percent (12%) of older patients presented with a geriatric syndrome as their sole symptom compared to 3% of younger patients (p=0.002).

Ancillary laboratory and imaging findings on presentation are shown in Table 3. The most common radiological findings at presentation for both the young and older groups were bilateral patchy opacities on chest x-ray.

**Table 2.**
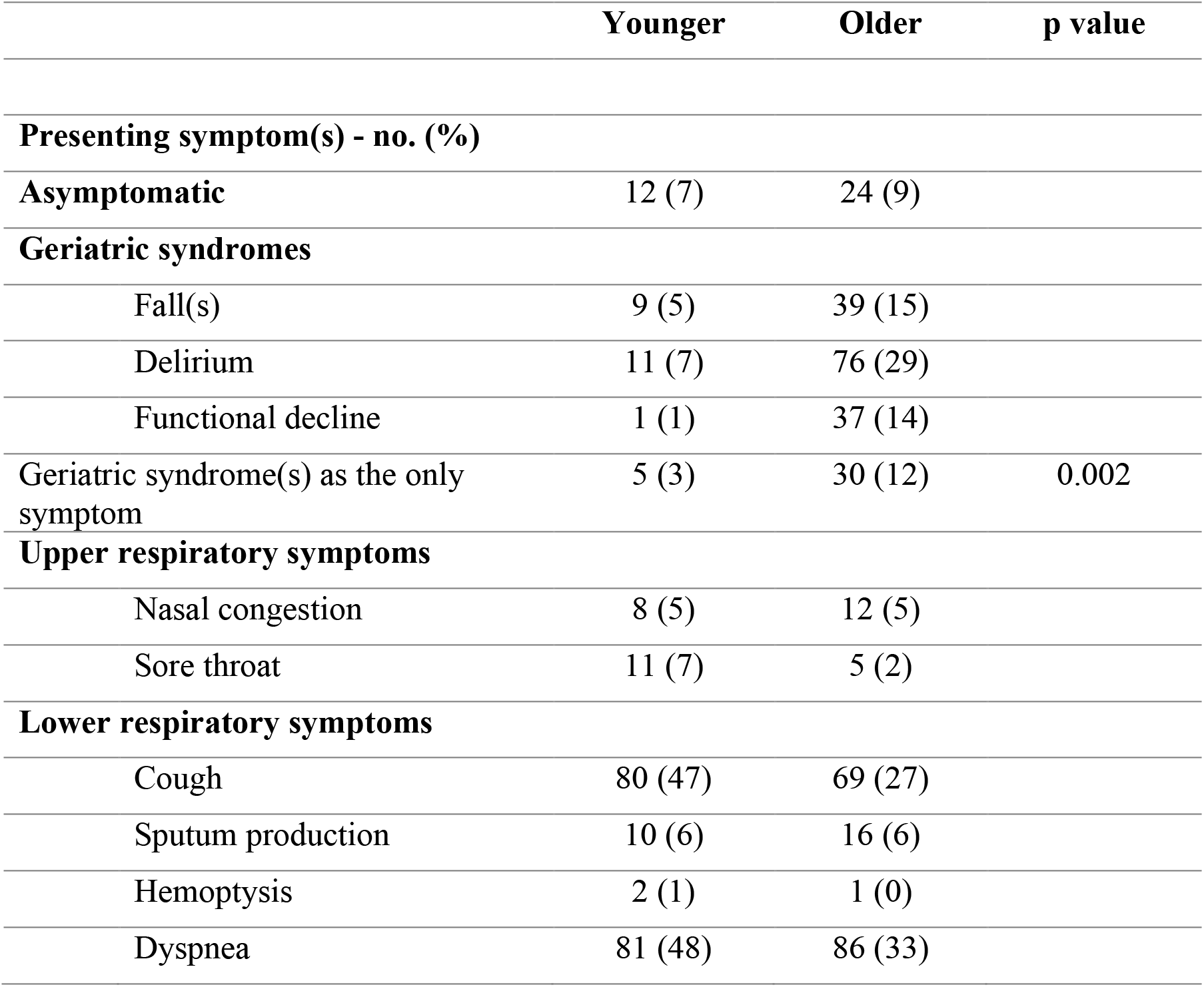
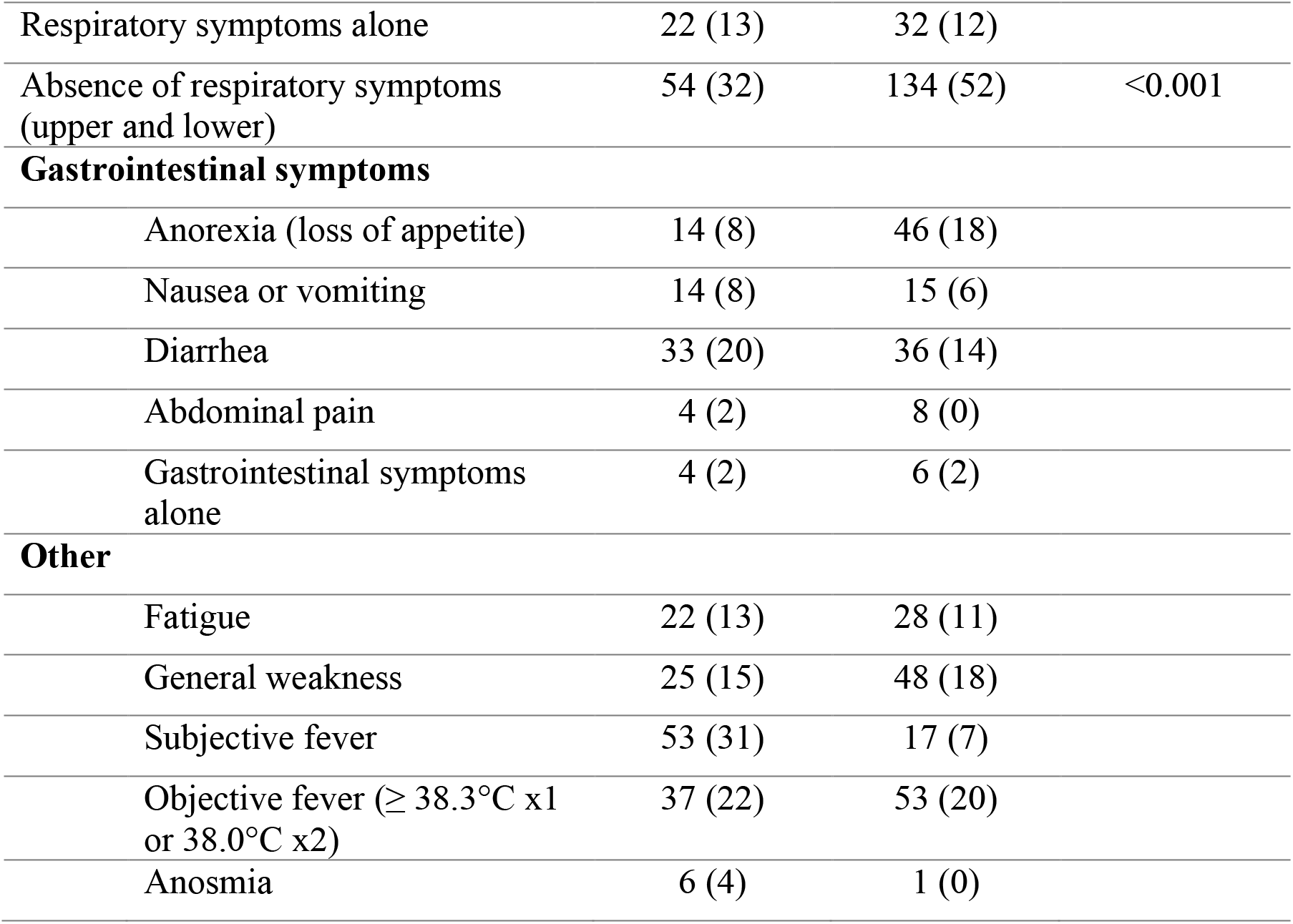
Clinical Presentation.

|  | Younger | Older | p value |
| --- | --- | --- | --- |
| **Presenting symptom(s) - no. (%)** |  |  |  |
| **Asymptomatic** | 12 (7) | 24 (9) |  |
| **Geriatric syndromes** |  |  |  |
| Fall(s) | 9 (5) | 39 (15) |  |
| Delirium | 11 (7) | 76 (29) |  |
| Functional decline | 1 (1) | 37 (14) |  |
| Geriatric syndrome(s) as the only symptom | 5 (3) | 30 (12) | 0.002 |
| **Upper respiratory symptoms** |  |  |  |
| Nasal congestion | 8 (5) | 12 (5) |  |
| Sore throat | 11 (7) | 5 (2) |  |
| **Lower respiratory symptoms** |  |  |  |
| Cough | 80 (47) | 69 (27) |  |
| Sputum production | 10 (6) | 16 (6) |  |
| Hemoptysis | 2 (1) | 1 (0) |  |
| Dyspnea | 81 (48) | 86 (33) |  |
| Respiratory symptoms alone | 22 (13) | 32 (12) |  |
| Absence of respiratory symptoms (upper and lower) | 54 (32) | 134 (52) | <0.001 |
| **Gastrointestinal symptoms** |  |  |  |
| Anorexia (loss of appetite) | 14 (8) | 46 (18) |  |
| Nausea or vomiting | 14 (8) | 15 (6) |  |
| Diarrhea | 33 (20) | 36 (14) |  |
| Abdominal pain | 4 (2) | 8 (0) |  |
| Gastrointestinal symptoms alone | 4 (2) | 6 (2) |  |
| **Other** |  |  |  |
| Fatigue | 22 (13) | 28 (11) |  |
| General weakness | 25 (15) | 48 (18) |  |
| Subjective fever | 53 (31) | 17 (7) |  |
| Objective fever ($\geq 38.3^{\circ}\text{C}$ x1 or $38.0^{\circ}\text{C}$ x2) | 37 (22) | 53 (20) | |
| Anosmia | 6 (4) | 1 (0) |  |

**Table 3.**
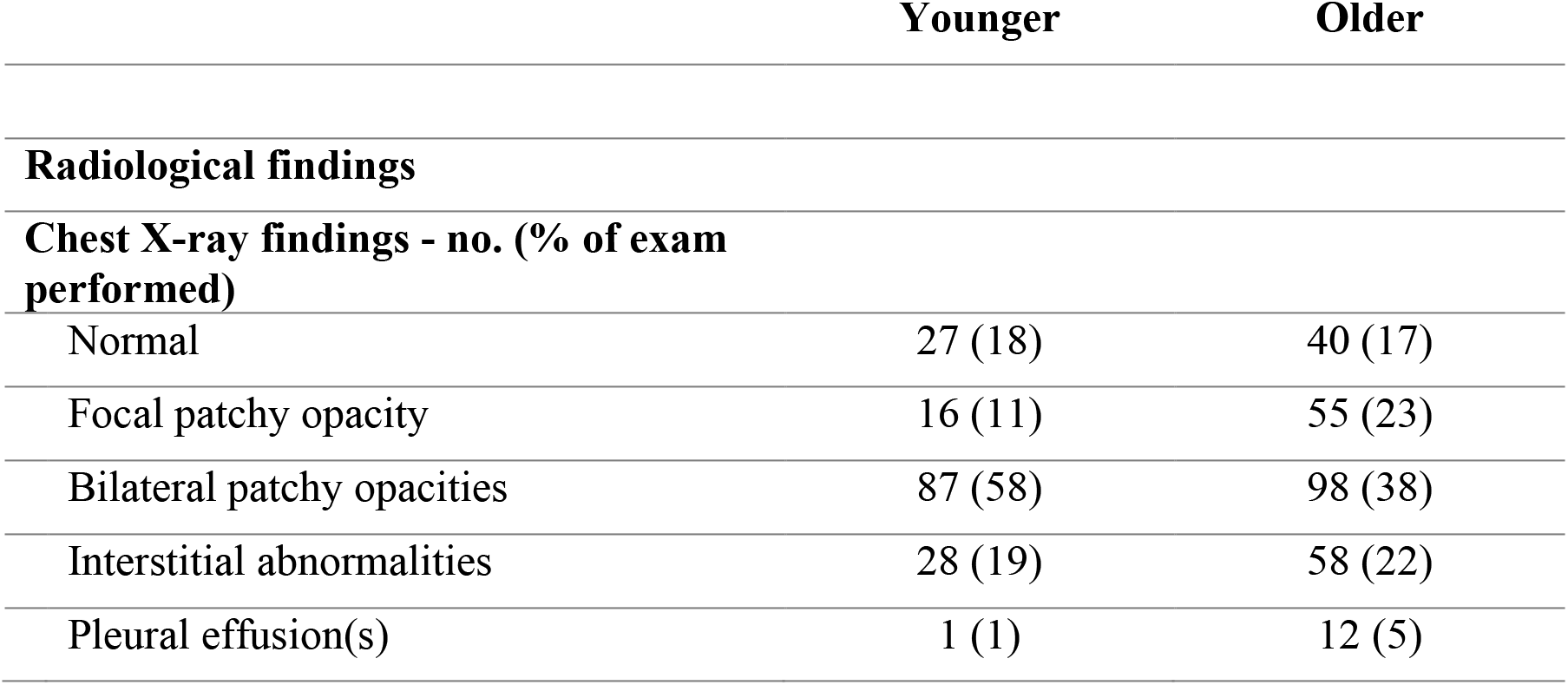
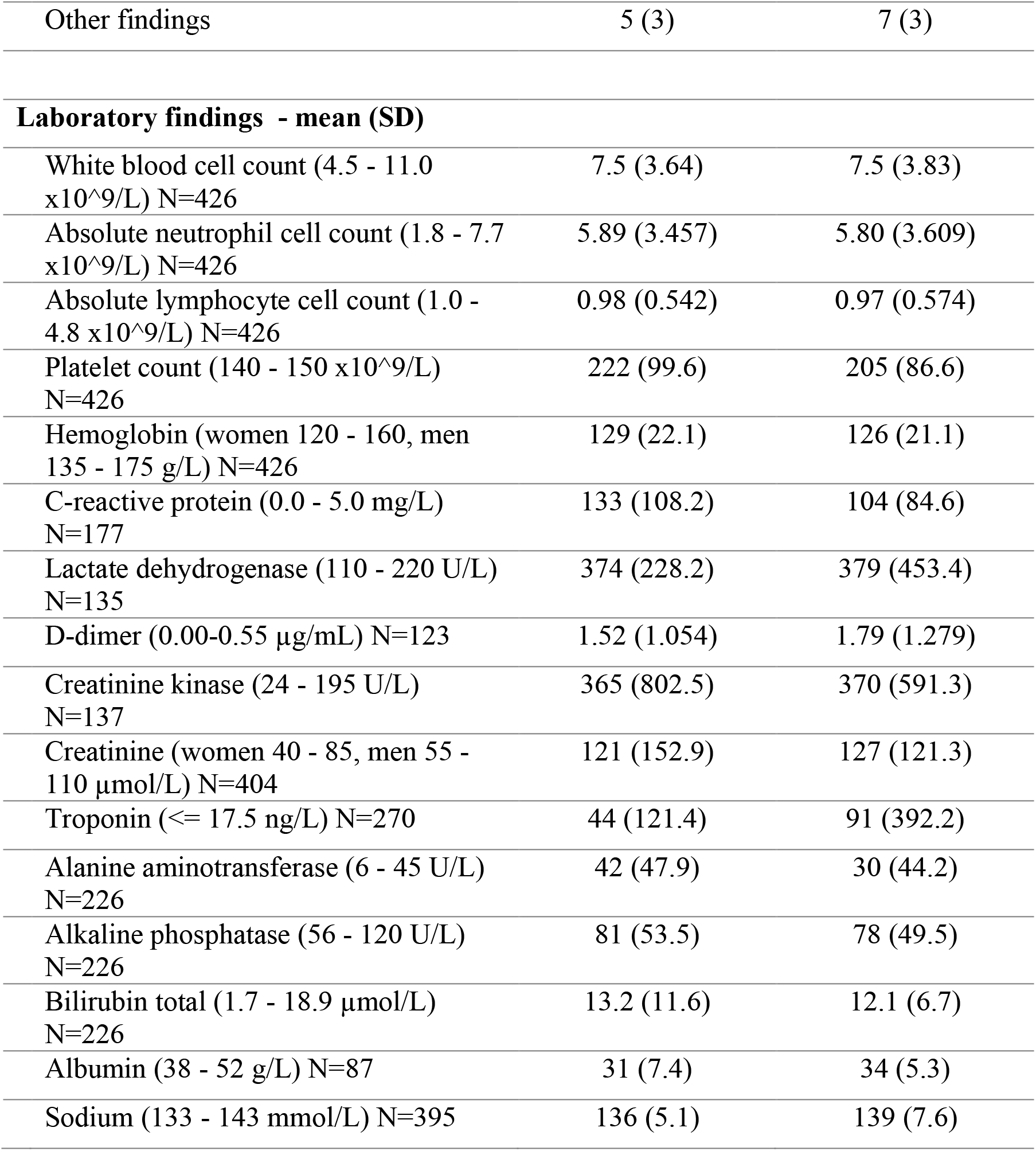
Ancillary Findings on Presentation.

|  | Younger | Older |
| --- | --- | --- |
| **Radiological findings** |  |  |
| **Chest X-ray findings - no. (% of exam performed)** |  |  |
| Normal | 27 (18) | 40 (17) |
| Focal patchy opacity | 16 (11) | 55 (23) |
| Bilateral patchy opacities | 87 (58) | 98 (38) |
| Interstitial abnormalities | 28 (19) | 58 (22) |
| Pleural effusion(s) | 1 (1) | 12 (5) |
| Other findings | 5 (3) | 7 (3) |
| **Laboratory findings - mean (SD)** |  |  |
| White blood cell count (4.5 - 11.0 x10<sup>9</sup>/L) N=426 | 7.5 (3.64) | 7.5 (3.83) |
| Absolute neutrophil cell count (1.8 - 7.7 x10<sup>9</sup>/L) N=426 | 5.89 (3.457) | 5.80 (3.609) |
| Absolute lymphocyte cell count (1.0 - 4.8 x10<sup>9</sup>/L) N=426 | 0.98 (0.542) | 0.97 (0.574) |
| Platelet count (140 - 150 x10<sup>9</sup>/L) N=426 | 222 (99.6) | 205 (86.6) |
| Hemoglobin (women 120 - 160, men 135 - 175 g/L) N=426 | 129 (22.1) | 126 (21.1) |
| C-reactive protein (0.0 - 5.0 mg/L) N=177 | 133 (108.2) | 104 (84.6) |
| Lactate dehydrogenase (110 - 220 U/L) N=135 | 374 (228.2) | 379 (453.4) |
| D-dimer (0.00-0.55 µg/mL) N=123 | 1.52 (1.054) | 1.79 (1.279) |
| Creatinine kinase (24 - 195 U/L) N=137 | 365 (802.5) | 370 (591.3) |
| Creatinine (women 40 - 85, men 55 - 110 µmol/L) N=404 | 121 (152.9) | 127 (121.3) |
| Troponin (<= 17.5 ng/L) N=270 | 44 (121.4) | 91 (392.2) |
| Alanine aminotransferase (6 - 45 U/L) N=226 | 42 (47.9) | 30 (44.2) |
| Alkaline phosphatase (56 - 120 U/L) N=226 | 81 (53.5) | 78 (49.5) |
| Bilirubin total (1.7 - 18.9 µmol/L) N=226 | 13.2 (11.6) | 12.1 (6.7) |
| Albumin (38 - 52 g/L) N=87 | 31 (7.4) | 34 (5.3) |
| Sodium (133 - 143 mmol/L) N=395 | 136 (5.1) | 139 (7.6) |

**Table 4.** Course in Hospital and Outcomes.

|  | Younger | Older | p value |
| --- | --- | --- | --- |
| **Complications - no. (%)** |  |  |  |
| None | 74 (44) | 40 (15) |  |
| **Geriatric syndromes** |  |  |  |
| Fall(s) | 3 (2) | 7 (3) |  |
| Delirium | 29 (17) | 77 (30) |  |
| Pressure injury | 3 (2) | 10 (4) |  |
| Malnutrition or denutrition | 6 (4) | 24 (9) |  |
| Deconditioning | 8 (5) | 21 (8) |  |
| **Cardiovascular** |  |  |  |
| NSTEMI or UA | 11 (7) | 34 (13) |  |
| STEMI | 12 (7) | 34 (13) |  |
| **Renal and metabolic** |  |  |  |
| AKI | 37 (22) | 92 (35) |  |
| Hyponatremia ($\leq 128$ mmol/L) | 22 (13) | 21 (8) | |
| Hypernatremia ($\geq 148$ mmol/L) | 28 (17) | 82 (32) | |
| Rhabdomyolysis | 1 (1) | 5 (2) |  |
| Transaminitis (ALT x3 UNL) | 21 (12) | 13 (5) |  |
| **Infectious** |  |  |  |
| Clostridioides difficile infection | 1 (1) | 3 (1) |  |
| Septic shock | 8 (5) | 6 (2) |  |
| Bacterial pneumonia superinfection | 30 (18) | 44 (17) |  |
| **Hematological** |  |  |  |
| DIC | 1 (1) | 2 (1) |  |
| Thrombocytopenia ($<100 \times 10^9$) | 19 (11) | 41 (16) | |
| VTE |  |  |  |
| PE* | 7 (4) | 5 (2) |  |
| DVT | 1 (1) | 3 (1) |  |

\*: definitive diagnose via CT-angio or V/Q scan at any point in time during admission

Intensive care and ventilatory support
|  |  |  |  |
| --- | --- | --- | --- |
| Admission to ICU if within goals of care (N=262) | 62 (43) | 35 (29) |  |
| Intubation and mechanical ventilation (N=262) | 36 (25) | 18 (15) |  |
| **Outcome (at date cutoff)** |  |  |  |
| Discharged from hospital - no. (%) | 148 (88) | 176 (67) |  |
| Mean length of stay - days (SD) | 19 (19.0) | 21 (14.6) |  |
| Death - no. (%) | 21 (12) | 84 (32) | <0.001 |
| Mean length of stay - days (SD) | 19 (17.3) | 16 (20.5) |  |

Both groups had lymphopenia on presentation (absolute lymphocyte count <1.0 x10^9^/L). Older patients had lower CRP compared to the younger group (mean 104 vs 133 mg/L). The older group had higher baseline albumin than the younger group (mean 31 vs 34 g/L).

### Complications and Outcomes

Fifteen percent (15%) of patients in the older group developed no complications from their disease compared to 44% in the younger group.

Older patients were more likely to develop delirium in hospital (30% vs 17%), non-ST segment elevation myocardial infarction (NSTEMI) (13% vs 7%), malnutrition (9% vs 4%), acute kidney injury (AKI) (35% vs 22%), and hypernatremia (32% vs 17%).

As per institutional policy, goals of care were established at the time of hospitalization. Two hundred sixty-two (262) of 429 patients were eligible for intensive care unit (ICU) admission. Twenty-nine percent (29%) of (? Eligible) older patients were cared for in the ICU, compared to 43% of younger patients.

In hospital mortality was higher in the older group (32% vs 12%, p<0.001).

## DISCUSSION

We report the characteristics, presenting features, and in-hospital outcomes of 429 COVID-19 affected patients hospitalized at the MUHC in Montreal, Canada, with particular attention to those above age 70. Patients ≥70 differed from younger patients in their presentations. Over half of older adults lacked any respiratory symptoms. Their non-respiratory presentations included geriatric syndromes of functional decline, fall, and delirium, which were the only symptoms in 12% of the older group. Older adults were more likely to develop delirium in hospital, cardiac complications, malnutrition, acute kidney injury, hypernatremia, and to die of their disease compared to patients younger than 70.

The high rate of non-respiratory COVID-19 presentations in our study is concordant with recent reports from France and the United Kingdom where delirium and falls were common reasons for hospitalizations in patients above age 80 with COVID-19 (8, 9, 12). Furthermore, given the high rate of patients from long-term care facilities in our study, our findings support the suggestion that non-respiratory presentations of COVID-19 are especially prominent in nursing home residents. This was shown in a single-center cohort study in the United Kingdom where 18.8% of patients from nursing homes had atypical symptoms, as well as in a point-prevalence survey in United States nursing homes where 8% of COVID-19 positive patients had atypical symptoms, defined in both studies as absence of fever, cough, or dyspnea (9, 13).

Our study does not reproduce the trend for elevated CRP and more profound lymphopenia in older adults on presentation compared to younger patients that was observed in China (14, 15). However, these studies use an age of 60 as cut-off to classify patients as elderly. Our results are more concordant with a study from the United Kingdom that showed no significant difference in inflammatory markers that used an age cut-off of 80 years old, which is closer to what would be defined as old age in Canada (9).

In addition, some older patients from our cohort were asymptomatic (9%) or mildly ill and hospitalized for isolation reasons, as they were living in collective-dwellings. This may explain 1) their less profound laboratory abnormalities and 2) their lower likelihood to be admitted to ICU if within goals of care. This contrasts with findings of a study from China were higher rates of ICU admission were observed in patients >60 (14).

Older adults in our study were more likely to develop acute kidney injury and hypernatremia in-hospital compared to their younger counterparts. A French study of 821 patients hospitalized to Geriatric wards with COVID-19 similarly observed AKI as a frequent complication, which was significantly more frequent in older adults who died (16). The occurrence of these complications suggests a vulnerability of older adults hospitalized with COVID-19 to dehydration. Dehydration in older persons can have severe consequences. If left untreated, mortality can exceed 50% (17, 18). Furthermore, it is a common precipitating factor for geriatric syndromes, including delirium and falls (19, 20). Cardiac complications were also more common in the older group. Of note, several of the NSTEMIs observed in our study population were classified by treating physicians as demand ischemia (type 2 NSTEMI) rather than plaque rupture, which may also suggest changes in volume status.

Our findings are consistent with previous studies that have demonstrated an association between older age and mortality (3, 7, 21). However, our study may underestimate complications and mortality in the elderly population as multiple patients were discharged to community inpatient centers in order to offload acute care beds during the first wave of the pandemic in Montreal. We suspect multiple patients may have died in the community.

### Limitations

Our study has limitations. First, the study only included patients within one center of the Montreal area (MUHC), but we included every patient, giving a comprehensive portrait in the most affected geographic area during the first wave of COVID-19 in Quebec. Second, the retrospective nature of the study required reliance on review of medical records for data extraction. Certain information was missing from baseline assessment, was based on other physicians’ interpretation of findings, or relied on patients’ recall of events. We only assessed short-term follow-up for all patients, limiting our assessment of long-term repercussions of the disease. Lastly, we performed statistical analyses for our pre-defined hypotheses regarding frequency of non-respiratory presentations, geriatric syndromes, and mortality. All other comparisons are descriptive. Caution should be taken regarding the reproducibility of the findings, with results viewed as hypothesis generating and to be validated in larger studies.

### Conclusion

This is the first study reporting clinical presentation, in-hospital course, and outcomes of elderly patients hospitalized with COVID-19 in Canada. The data emphasizes the importance of non-respiratory presentations, and especially of geriatric syndromes as presenting features of COVID-19 in older adults. Our findings further demonstrate the heightened vulnerability of older adults to complications related to dehydration in hospital, emphasizing the need for early well-coordinated multidisciplinary care upon admission for this population.

### Implications for Practice

- Beware of non-respiratory presentation of COVID-19 in older persons. An older person developing a new geriatric syndrome, in the context of the COVID-19 pandemic, should prompt early isolation and testing.
- When caring for older adults hospitalized with COVID-19, pay specific attention to evaluating and supporting proper hydration. Monitor closely for cardiac complications.

## Data Availability

Data referred to in this article has been anonymized and is kept with the study investigator.

